# The effects of vortioxetine versus placebo on cognitive functioning in mild cognitive impairment: a placebo-controlled, randomized, double-blind study

**DOI:** 10.64898/2025.12.10.25341977

**Authors:** V Matev, M Rothenberg, J Donath, C Cukaci, M Klöbl, K Al Barede, P Stöhrmann, G Dörl, R Lanzenberger, D Winkler, E Winkler-Pjrek

## Abstract

**Introduction:** This study investigates the effects of vortioxetine on cognitive performance in patients with mild cognitive impairment (MCI) in a randomized controlled trial (RCT). MCI is characterized by memory impairment and deficits in other cognitive domains and has been found to progress into Alzheimer’s dementia (AD) in some cases. In a recent open-label, single-arm study, vortioxetine, an antidepressant, was associated with improved cognitive function in patients with MCI, however, no placebo-controlled studies have been conducted to support these findings.

**Methods:** Forty-seven subjects diagnosed with MCI were randomly assigned to receive treatment with either 10 mg vortioxetine, 20 mg vortioxetine, or placebo over 12 weeks, in a randomized, double-blind study design. Neuropsychological tests were conducted several times during the trial, including at a baseline screening visit, consisting of the Alzheimer’s Disease Assessment Scale – Cognitive Subscale (ADAS-cog), a German version of the Rey Auditory Verbal Learning Test (“Verbaler Lern-und Merkfähigkeitstest”, VLMT), the Digit Symbol Substitution Test (DSST), the Mini-Mental State Examination (MMSE), and the Quality of Life Scale (QOLS). Statistical analysis of neuropsychological data was performed using linear mixed models (LMM) and subsequent post-hoc testing to account for multiple comparisons.

**Results:** LMM results showed a significant increase in VLMT test scores from baseline to week 12 in the 10 mg (p = 0.02) and 20 mg vortioxetine groups (p = 0.017). The placebo group showed a significant increase in test scores from baseline to week 4 (p = 0.009), followed by a significant decrease in test performance observed from week 4 to week 8 (p = 0.006). Both the vortioxetine and placebo groups showed statistically significant DSST test score changes over 12 weeks (10mg: p = 0.004; 20mg: p <.0001; placebo: p <.0001). Similarly, ADAS-Cog test performance significantly increased from baseline to week 12 in both the 10 mg vortioxetine group (p = 0.02) and the placebo group (p = 0.002). MMSE test results showed significant improvement over 8 weeks in the 20 mg vortioxetine group (p = 0.011), compared to no significant findings in the 10 mg and placebo groups. A significant improvement of QOLS scores was observed from week 8 to week 12 in the 10 mg vortioxetine group only (p = 0.033).

**Conclusion:** The results of this study indicate a potential treatment effect of vortioxetine on verbal learning and memory processes in patients with MCI. However, the effects of vortioxetine could not be found at the group level; thus, further research and larger RCTs are needed to elucidate its underlying mechanisms and potential in MCI and early AD treatment.

## INTRODUCTION

Mild cognitive impairment (MCI) is a syndrome characterized by memory impairment and other cognitive deficits beyond that expected for age and education (Petersen et al, 1999) and is considered a prodromal stage for dementia (Langa & Levine, 2014). The overall prevalence of MCI in the general population has been estimated between 12% and 18% in persons above 60 years (Petersen, 2016), with an annual conversion rate to Alzheimer’s Disease (AD) between 5% and 10% compared to 1% to 2% per year in healthy adults (Grundman et al, 2004). Alzheimer’s disease – a chronic and progressive neurodegenerative disease – is thought to be the most common cause of dementia, with about 50 million people affected worldwide and a yearly incidence of 10 million, as estimated by the World Health Organization (WHO, 2025). Due to increases in life expectancy, these numbers are expected to increase further during the next 30 years (Alzheimer’s Association, 2016). Patients with MCI and AD face a progressive cognitive impairment, which not only results in a deterioration of functional independence and quality of life, but is also associated with high costs due to the functional decline of patients and subsequent need for nursing care in more advanced disease stages (Callahan, 2017).

Neuroimaging studies have discovered alterations in resting-state functional connectivity (FC) in AD (Damoiseaux, 2012) and MCI (Sheline & Raichle, 2013), specifically loss of intranetwork connectivity in large-scale brain networks, such as the default mode network, the control network, the dorsal attention network, the salience network, and the sensory motor network. Additionally, between-network connectivity seems to deteriorate as the disease progresses (Sheline & Raichle, 2013).

Furthermore, consistent evidence has been found regarding the degeneration of monoaminergic systems, particularly the serotonergic neurotransmitter system, in AD (Lanctot at al, 2007). In MCI, similar mechanisms have been discovered. Studies evaluating the pathogenesis of MCI have reported a degeneration of the serotonergic system in MCI patients (Hasselbach et a, 2008; Smith et al, 2017), with evidence suggesting a more pronounced effect of serotonin (5-HT) transporter binding reductions than grey matter atrophy in MCI (Smith et al, 2017). These serotonergic alterations, specifically the decreased 5-HT transporter availability in the dorsal raphe nucleus, seem to be related to altered FC of hippocampal regions (Barrett et al, 2017). This link may offer a neurobiological explanation for memory impairments in MCI and AD, since serotonergic projections from the raphe nuclei to the hippocampus play an essential role in supporting neurogenesis in the hippocampus (Alenina & Klempin, 2015).

The treatment of cognitive symptoms in AD comprises symptomatic pharmacotherapy with acetylcholinesterase (AChE) inhibitors, memantine, a N-methyl-D-aspartate receptor antagonist, antibody treatments, such as donanemab and lecanemab, as well as a range of non-pharmacological interventions including cognitive training, physical exercise, and occupational therapy (Szeto & Lewis, 2016; Chen et al, 2025). A meta-analysis by Dou, et al. (2018) has found that galantamine and donepezil are most effective for mild to moderate AD, whereas a combination of memantine and donepezil has been shown to alleviate symptomatology in moderate to severe AD, with an effect size of 0.76 (Devita et al, 2021). Nevertheless, treatment with memantine has not been proven effective for MCI (Schneider et al., 2011). Similarly, a meta-analysis comparing 14 RCTs on cholinesterase inhibitors for the treatment of MCI has found no differences in cognitive functioning when compared to placebo, although the use of cholinesterase inhibitors was associated with a lower turnover rate from MCI to AD (Matsunaga et al., 2019). Since 2023, phase 3 trials for lecanemab and donanemab, agents targeting the underlying neuropathology of AD, have been completed, paving the way for disease-modifying treatment options in neurodegenerative diseases such as AD (Haneka et al., 2024). The mechanism of action of these humanized IgG-1 monoclonal antibodies primarily involves targeting aggregated β-amyloid species, specifically β-amyloid 42 protofibrils (lecanemab) and N-terminal pyroglutamate β-amyloid epitopes that are present in established plaques (donanemab) (Haneka et al., 2024). However, many of the now available treatment options for AD and MCI lack conclusive evidence of efficacy and safety profiles, as anti-β amyloid monoclonal antibodies may lead to unfavorable side effects like APOE-dependent local brain oedema and bleeding (ARIA-E and-H) or infusion-related reactions (Fox et al., 2025). Furthermore, possible interaction effects with other commonly prescribed drugs increase hinder a number of patients to receive these treatments, increasing the need for alternative treatment options for these patients.

As serotonergic compounds such as paroxetine have also been shown to be able to reduce amyloid beta peptide in the cortex of a transgenic mouse model of AD (Nelson et al, 2007), the serotonergic system might be an interesting target for prevention and early treatment of MCI patients. In a recent open-label, single-arm study, vortioxetine, a novel antidepressant acting on the serotonergic pathways, was associated with improved cognitive function in MCI patients without depression (Tan & Tan, 2021). Vortioxetine has been licensed for the treatment of major depressive disorder since 2013 (D’Agostino, et al, 2015), with further clinical data supporting its use in anxiety disorders (Yee et al, 2018). The substance binds to six molecular target sites, by inhibiting the 5-HT transporter, acting as an agonist at the 5-HT1A receptor, as well as a partial agonist at the 5-HT1B receptor and as antagonist at the 5-HT1D, 5-HT3, and 5-HT7 receptors (Sanchez et al, 2015). Furthermore, vortioxetine has been shown to increase the release of norepinephrine, dopamine, acetylcholine, and histamine (Mork et al, 2012), possibly explaining its effects on cognitive function. Several studies have found improvements in executive function, attention, processing speed, and memory in patients with major depressive disorder treated with vortioxetine (Mahableshwarkar et al, 2015; McIntyre et al, 2014; McIntyre et al, 2017; Harrison et al, 2016). Furthermore, improvements in neuropsychological tests along with changes in fMRI BOLD signal have also been observed in remitted depressed patients and healthy volunteers (Smith et al, 2018). The efficacy of vortioxetine on cognitive function in major depression seems to be a unique effect when compared to other antidepressants commonly used to treat major depressive disorder, as a recent meta-analysis showed that vortioxetine was the only substance that improved cognitive dysfunction on the Digit Symbol Substitution Test vs. placebo (Baune et al, 2018). Vortioxetine appears to be safe and well-tolerated, both in short-and long-term treatment. The rate of sexual dysfunction, insomnia, weight gain and discontinuation symptoms seem to occur with a lower incidence than in other serotonergic and noradrenergic antidepressants (Baldwin et al, 2016). Taken together, the observation of an early serotonergic dysfunction in the pathogenesis of AD and the positive cognitive effects seen with vortioxetine treatment might render this drug a promising option for symptomatic or even disease-modifying treatment in MCI. However, clinical data on the use of vortioxetine in MCI and AD is lacking. To date, only one study has assessed the effects of vortioxetine on cognitive function in patients diagnosed with MCI, showing that vortioxetine was associated with significantly improved MoCA-, DSST-, CIBIC+-and global CDR scores (Tan & Tan, 2021). These findings are in line with previous research in MDD patients, indicating an independent effect of vortioxetine on cognitive performance with significant improvements in DSST scores compared to placebo (Katona et al., 2012; McIntyre et al., 2014; Mahableshwarkar et al., 2015). However, the role of vortioxetine in the treatment of MCI, along with the specific pathophysiological implications of this treatment, has not been investigated in a placebo-controlled randomized controlled trial before. Thus, this study aims to investigate the effects of vortioxetine on cognitive performance in patients with MCI, expecting a dose-dependent statistically significant improvement of neuropsychological test scores with vortioxetine compared to placebo.

## MATERIALS AND METHODS

Subjects diagnosed with MCI, without depressive symptoms, were randomly assigned to receive treatment with either 10mg vortioxetine, 20mg vortioxetine, or placebo over 12 weeks, in a double-blind study design. The study was conducted between 2021 and 2024 at the Medical University of Vienna, enrolling a total of 47 participants. Subjects were recruited at the memory clinic of the Department of Psychiatry and Psychotherapy at the Medical University of Vienna, as well as through advertisements in local newspapers. Neuropsychological tests were conducted several times during the trial, including at baseline before treatment (Week-1). All subjects provided written informed consent before study entry. The independent ethics committee of the Medical University of Vienna approved the study protocol and all related forms and amendments of this study (ethical committee number 1491/2019). The clinical trial was performed in full compliance with the legal regulations according to the Austrian Drug Law (AMG - Arzneimittelgesetz) and was registered in the European Clinical Trial Database (EudraCT 2019-001836-69).

### Participants

Study subjects were 54 to 80 years of age (mean age 70 ± 6.86, 31 male, 16 female) and met criteria for MCI following the criteria stipulated by the National Institute on Aging – Alzheimer’s Association (NIA-AA) workgroups on diagnostic guidelines for Alzheimer’s disease (Albert et al, 2011). Subjects meeting any of the following criteria during screening or during the following study visits were excluded from the study: Previous or current DSM-5 axis-I disorder, diagnosis of dementia, evidence of previous or current significant neurological disorder, use of prohibited concomitant medication such as antidepressants, antipsychotics, antidementia medication and anticonvulsant medication, pacemakers, cochlear implants, insulin pumps or other implants made of ferromagnetic materials, or metal splinters in the body.

### Study Medication

After the initial screening visit (Week-1), subjects were randomly assigned to one of the three treatment arms, vortioxetine 10mg, 20mg, or placebo, using simple randomization (1:1:1 ratio) at visit 2 (Week 0). Randomization was carried out by a designated member of the research group not involved in the clinical treatment of the study subjects, using a computer program. Treatment with vortioxetine was started at a daily dose of 5mg for 3 days. Patients in the vortioxetine 20mg group received 10mg for an additional 3 days before the dose increase to 20mg. Tablets were taken orally once a day in the morning for a total duration of 12 weeks, as prior studies with vortioxetine in depressed patients have found significant improvements in cognition within 8 weeks (McIntyre et al, 2014), as well as studies in Alzheimer’s dementia showing significant cognitive improvements with acetylcholinesterase inhibitors after 8 and 12 weeks of treatment (Gauthier et al, 2002). Compliance was assessed by patient reports and pill counts of the returned study medication.

### Medical and Psychological Assessments

Relevant medical, neurological, and psychiatric history, current medical and psychiatric conditions, and current medications were recorded prior to the start of treatment. Subjects were screened via a complete physical, neurological, and psychiatric examination by an experienced psychiatrist or psychiatrist in training under the supervision of a senior psychiatrist to provide information on their symptomatology and clinical status. Neuroimaging (resting state-fMRI and T1-weighted, structural MRI) was carried out before inclusion in order to adequately assess AD-typical atrophies in the brain as well as significant vascular lesions that would lead to an exclusion of study participation.

Psychiatric evaluation was based on the Structured Clinical Interview for DSM-5 (SCID-5) (First et al, 2015) as well as a screening for acute suicidality. Clinical examination included ECG, blood pressure measurements, and blood/urine tests. In addition, subjects were screened for reversible causes of memory impairment. Baseline laboratory tests comprised haematology, biochemistry, coagulation, thyroid hormones, vitamin B12, folic acid, glycated hemoglobin, serological tests for syphilis and HIV, parathyroid hormone, and urine drug screening test. Follow-up laboratory tests at visits 5 and 7 included hematology, clinical chemistry, coagulation, and thyroid hormones.

Neuropsychological tests for the assessment of cognitive functioning, as well as psychological questionnaires, were conducted at several time points throughout this study, consisting of the Alzheimer’s Disease Assessment Scale – Cognitive Subscale (ADAS-cog) (Rosen et al, 1984), a German version of the Rey Auditory Verbal Learning Test (Lezak et al, 2012) (“Verbaler Lern-und Merkfähigkeitstest”, VLMT (Helmstaedter et al, 2001)), the Digit Symbol Substitution Test (DSST) (Jaeger, 2018), the Mini-Mental State Examination (MMSE) (Folstein et al, 1983), the Clinical Global Impression of Severity (CGI-S), the Clinical Global Impression of Improvement (CGI-I), the CGI Efficacy Index (CGI-EI) (Guy, 1976), the Quality of Life Scale (QOLS) (Offenbacher et al, 2012), and the Geriatric Depression Scale (GDS) (Yesavage et al, 1982). To minimize learning effects, three parallel versions of the VLMT were used and alternated through visits 1, 5, 6, and 7. The tolerability of vortioxetine was systematically assessed during the drug trial with the Udvalg for Kliniske Undersogelser Side Effect Rating Scale (UKU scale) (Lingjaerde et al, 1987).

### Statistical Analysis

Sample size considerations were based on a meta-analysis evaluating twelve short-term (6 to 12 weeks) randomized controlled trials on vortioxetine, reporting an average Cohen’s d of 0.217 (f=0.11) for the antidepressant efficacy compared to placebo (Pae et al., 2015). Based on an α-error of 0.05 per test and around 300 parcels for connectivity analysis (Eickhoff et al, 2018), a corrected α-error probability of 1.71E-4 (Sidak-adjusted) at a power of 0.8 and moderate correlation between repetitions of r=0.5 yielded an overall sample size of N=36. Using 15 subjects per group increased the power to 0.95, allowing for non-sphericity corrections, compensating for lower correlations between repetitions, or allowing for considerably more tests (>8000).

Statistical analysis of neuropsychological data was performed using the R Project for Statistical Computing (https://www.r-project.org/). Changes in cognitive performance, measured by total test scores, were analyzed using linear mixed models (LMM), with time as a within-subjects factor and treatment group (vortioxetine 10mg, 20mg, or placebo) as a between-subjects factor, with the significance level set at α = 0.05. Post-hoc Tukey tests were computed to account for multiple comparisons. For non-normal distributed data, evaluated by the Shapiro-Wilk test, a Friedman Test for within-subject factors and a Kruskal-Wallis test for between-subject factors were calculated instead, using post-hoc Wilcoxon Tests (Bonferroni-corrected) to further evaluate significant results.

**Figure 1.**
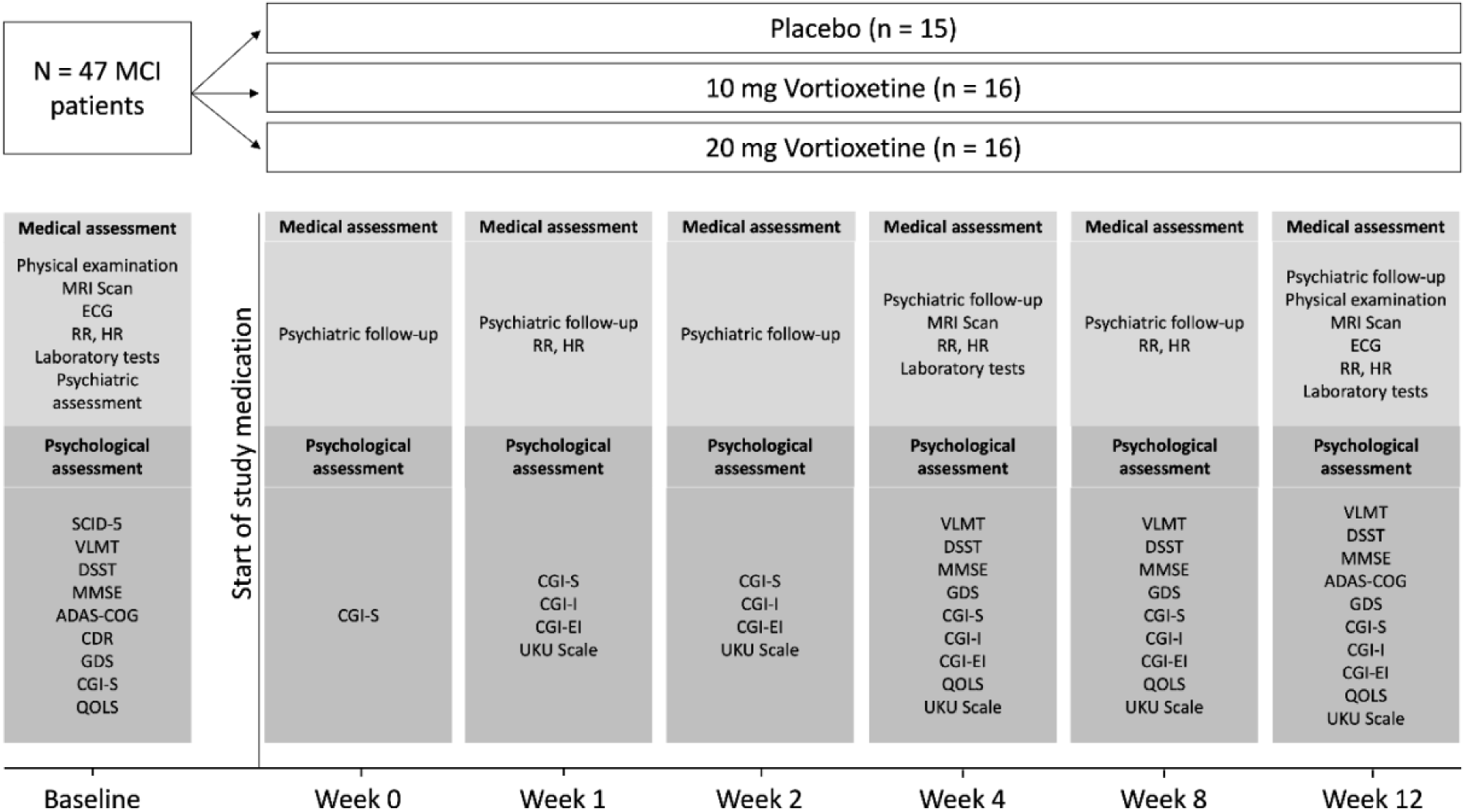
Study design: 47 subjects were randomly assigned to either one of three treatment arms (Placebo, Vortioxetine 10mg or Vortioxetine 20mg). At baseline and at several timepoints during the study, medical and psychological assessments were conducted. The results of the neuroimaging findings during this trial will be published in a separate article.

## RESULTS

A total of 47 MCI patients were randomized to either one of the three treatment arms (vortioxetine 10 mg, vortioxetine 20 mg or placebo). Baseline demographics were similar across all groups (Table 1).

**Table 1.** Baseline Demographics and Test Scores.

|  | Vortioxetine 10 mg<br>(n = 16) | Vortioxetine 20 mg<br>(n = 16) | Placebo<br>(n = 15) |
| --- | --- | --- | --- |
| *Age (years)* |  |  |  |
| Mean (SD) | 70.7 (8.03) | 71.2 (6.96) | 72.5 (5.06) |
| Median (min, max) | 72.7 (56.0, 79.5) | 70.6 (54.9, 80.3) | 73.1 (62.6, 79.0) |
| *Sex* |  |  |  |
| Male | 10 (62.5%) | 9 (56.2%) | 11 (73.7%) |
| Female | 8 (37.5%) | 7 (43.8%) | 4 (26.7%) |
| *VLMT* |  |  |  |
| Mean (SD) | 43.9 (11.2) | 44.0 (12.2) | 41.7 (11.3) |
| *DSST* |  |  |  |
| Mean (SD) | 43.8 (11) | 38.9 (10.4) | 38.3 (13.4) |
| *ADAS-Cog* |  |  |  |
| Mean (SD) | 14.5 (9.3) | 12.5 (5) | 12.7 (6.2) |
| *MMSE* |  |  |  |
| Mean (SD) | 28.3 (1.2) | 27.1 (2.7) | 27.7 (1.8) |
| *GDS* |  |  |  |
| Mean (SD) | 3.1 (3.1) | 2.7 (1.8) | 2.7 (2.3) |
| *QOLS* |  |  |  |
| Mean (SD) | 90.1 (13.7) | 90.4 (11.3) | 93.1 (12.6) |
| *CGI-S* |  |  |  |
| Mean (SD) | 2.6 (1.1) | 2.8 (1) | 3.0 (0.8) |

**Table 2.**
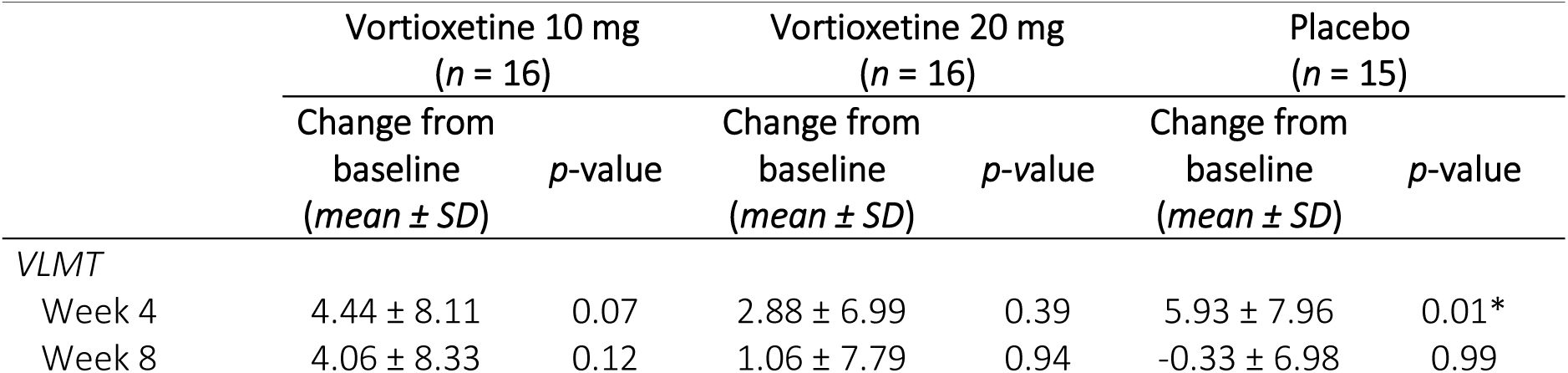
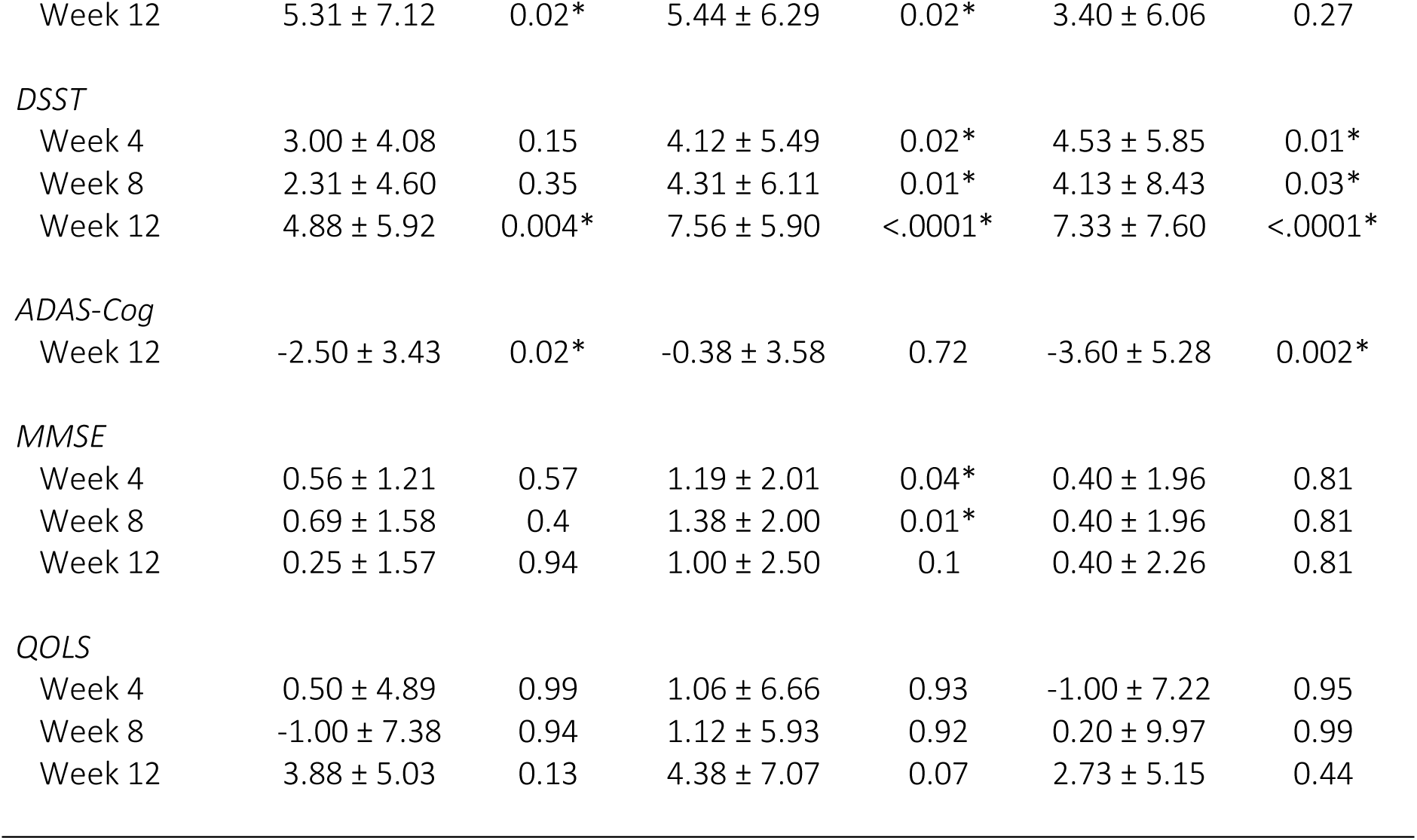
(Neuro)psychological Test Results.

|  | Vortioxetine 10 mg<br>(n = 16) |  | Vortioxetine 20 mg<br>(n = 16) |  | Placebo<br>(n = 15) |  |
| --- | --- | --- | --- | --- | --- | --- |
| | Change from<br>baseline<br>(mean ± SD) | p-value | Change from<br>baseline<br>(mean ± SD) | p-value | Change from<br>baseline<br>(mean ± SD) | p-value |
| *VLMT* |  |  |  |  |  |  |
| Week 4 | 4.44 ± 8.11 | 0.07 | 2.88 ± 6.99 | 0.39 | 5.93 ± 7.96 | 0.01* |
| Week 8 | 4.06 ± 8.33 | 0.12 | 1.06 ± 7.79 | 0.94 | -0.33 ± 6.98 | 0.99 |
| Week 12 | 5.31 ± 7.12 | 0.02* | 5.44 ± 6.29 | 0.02* | 3.40 ± 6.06 | 0.27 |
| *DSST* |  |  |  |  |  |  |
| Week 4 | 3.00 ± 4.08 | 0.15 | 4.12 ± 5.49 | 0.02* | 4.53 ± 5.85 | 0.01* |
| Week 8 | 2.31 ± 4.60 | 0.35 | 4.31 ± 6.11 | 0.01* | 4.13 ± 8.43 | 0.03* |
| Week 12 | 4.88 ± 5.92 | 0.004* | 7.56 ± 5.90 | <.0001* | 7.33 ± 7.60 | <.0001* |
| *ADAS-Cog* |  |  |  |  |  |  |
| Week 12 | -2.50 ± 3.43 | 0.02* | -0.38 ± 3.58 | 0.72 | -3.60 ± 5.28 | 0.002* |
| *MMSE* |  |  |  |  |  |  |
| Week 4 | 0.56 ± 1.21 | 0.57 | 1.19 ± 2.01 | 0.04* | 0.40 ± 1.96 | 0.81 |
| Week 8 | 0.69 ± 1.58 | 0.4 | 1.38 ± 2.00 | 0.01* | 0.40 ± 1.96 | 0.81 |
| Week 12 | 0.25 ± 1.57 | 0.94 | 1.00 ± 2.50 | 0.1 | 0.40 ± 2.26 | 0.81 |
| *QOLS* |  |  |  |  |  |  |
| Week 4 | 0.50 ± 4.89 | 0.99 | 1.06 ± 6.66 | 0.93 | -1.00 ± 7.22 | 0.95 |
| Week 8 | -1.00 ± 7.38 | 0.94 | 1.12 ± 5.93 | 0.92 | 0.20 ± 9.97 | 0.99 |
| Week 12 | 3.88 ± 5.03 | 0.13 | 4.38 ± 7.07 | 0.07 | 2.73 ± 5.15 | 0.44 |

**Table 3.** CGI Scores (*mean ± SD*)

|  | Vortioxetine 10 mg<br>(*n* = 16) | Vortioxetine 20 mg<br>(*n* = 16) | Placebo<br>(*n* = 15) |
| --- | --- | --- | --- |
| *CGI-S\** |  |  |  |
| Week -1 | 2.63 ± 1.09 | 2.75 ± 1 | 3 ± 0-76 |
| Week 0 | 2.5 ± 1.15 | 2.25 ± 1 | 3.27 ± 0.96 |
| Week 1 | 2.44 ± 1.15 | 2.25 ± 1 | 3.33 ± 1.18 |
| Week 2 | 2.63 ± 1.26 | 2.31 ± 1.02 | 3.33 ± 1.11 |
| Week 4 | 2.44 ± 1.03 | 2.06 ± 0.85 | 3.27 ± 1.28 |
| Week 8 | 2.56 ± 1.03 | 2.44 ± 1.31 | 3.07 ± 1.16 |
| Week 12 | 2.63 ± 1.26 | 2.19 ± 1.33 | 2.87 ± 1.3 |
| *CGI-I\*\** |  |  |  |
| Week 1 | 3.94 ± 0.25 | 3.81 ± 0.4 | 3.93 ± 0.26 |
| Week 2 | 3.94 ± 0.44 | 3.69 ± 0.48 | 3.93 ± 0.26 |
| Week 4 | 3.69 ± 0.6 | 3.56 ± 0.51 | 3.6 ± 0.5 |
| Week 8 | 3.56 ± 0.81 | 3.44 ± 0.73 | 3.67 ± 0.62 |
| Week 12 | 3.38 ± 0.89 | 3.5 ± 0.82 | 3.53 ± 0.64 |
| *CGI-EI* |  |  |  |
| Week 1 | 13.13 ± 1.26 | 12.31 ± 1.99 | 12.73 ± 1.03 |
| Week 2 | 12.81 ± 1.68 | 12.06 ± 1.91 | 12.73 ± 1.03 |
| Week 4 | 11.56 ± 2.9 | 11.31 ± 2.87 | 11.47 ± 1.96 |
| Week 8 | 11 ± 3.2 | 11 ± 2.68 | 11.8 ± 1.93 |
| Week 12 | 10.31 ± 3 | 11 ± 3.08 | 10.87 ± 2.97 |

\**CGI-S (Clinical Global Impression – Improvement)*: 0 Not assessed; 1 Normal, not at all ill; 2 Borderline mentally ill; 3 Mildly ill; 4 Moderately ill; 5 Markedly ill; 6 Severely ill; 7 Among the most extremely ill patients
\*\**CGI-I (Clinical Global Impression – Severity of Illness)*: 0 Not assessed; 1 Very much improved; 2 Much improved; 3 Minimally improved; 4 No change; 5 Minimally worse; 6 Much worse; 7 Very much worse

### Neuropsychological Test Scores

The mean change from baseline (week-1) to week 12 in VLMT performance score was 6.1 for the low dose vortioxetine group (10 mg), 5.6 for the higher dose group (20mg vortioxetine), and 3.1 for placebo. LMM results showed a significant increase of test scores over time and were confirmed by Tukey corrected post-hoc tests for baseline to week 12 in the 10mg (p = 0.02) as well as the 20mg vortioxetine group (p = 0.017). The placebo group showed a significant increase of test scores from baseline to week 4 (p = 0.009), followed by a significant decrease in test performance observed from week 4 to week 8 (p = 0.006). No significant effects between treatment groups could be observed.

DSST test score changes showed a mean increase of 6 points from week-1 to week 12 for the 10mg vortioxetine group, 7.9 points for the 20mg vortioxetine group, and 8.4 points for the placebo group. Both vortioxetine and the placebo groups produced statistically significant test score changes over a 12-week timespan (p = 0.004; p <.0001; p <.0001). Similarly, ADAS-Cog test performance significantly increased from baseline to week 12 in both the 10 mg vortioxetine group (p = 0.02) and the placebo group (p = 0.002).

MMSE test results showed significant improvement over the course of 8 weeks in the 20 mg vortioxetine group (p = 0.011), but no significant changes across the full study duration of 12 weeks in either of the treatment groups, nor at the group level when comparing vortioxetine vs. placebo.

### Secondary Outcome Measures

There was an average increase of 4.4 points in QOLS scores from visit 1 to visit 7 in the 10 mg vortioxetine group, 4.8 points in the 20 mg vortioxetine group, and 3.2 points in the placebo group, with a significant improvement of QOLS scores observed from week 8 to week 12 only in the 10 mg vortioxetine group (p = 0.033). GDS scores decreased on average by 0.2 points in the 10 mg vortioxetine group, 0.7 points in the 20 mg group, and 1.2 points in the placebo group. CGI-S scores produced statistically significant treatment group x time differences between the 20 mg vortioxetine group and placebo (p = 0.04), with significantly lower severity of illness scores in the 20 mg group up to week 4. CGI-I scores improved on average by 0.6 points in the 10 mg vortioxetine group, 0.31 points in the 20 mg group and 0.4 points in the placebo group, reaching significance only in the 10 mg vortioxetine group (week 1 to week 12, p = 0.006). Similarly, the efficacy index, calculated through CGI-EI scores, improved significantly in the 10 mg vortioxetine group (week 1 to week 12, p = 0.0004), compared to the 20 mg and placebo groups.

**Figure 2.**
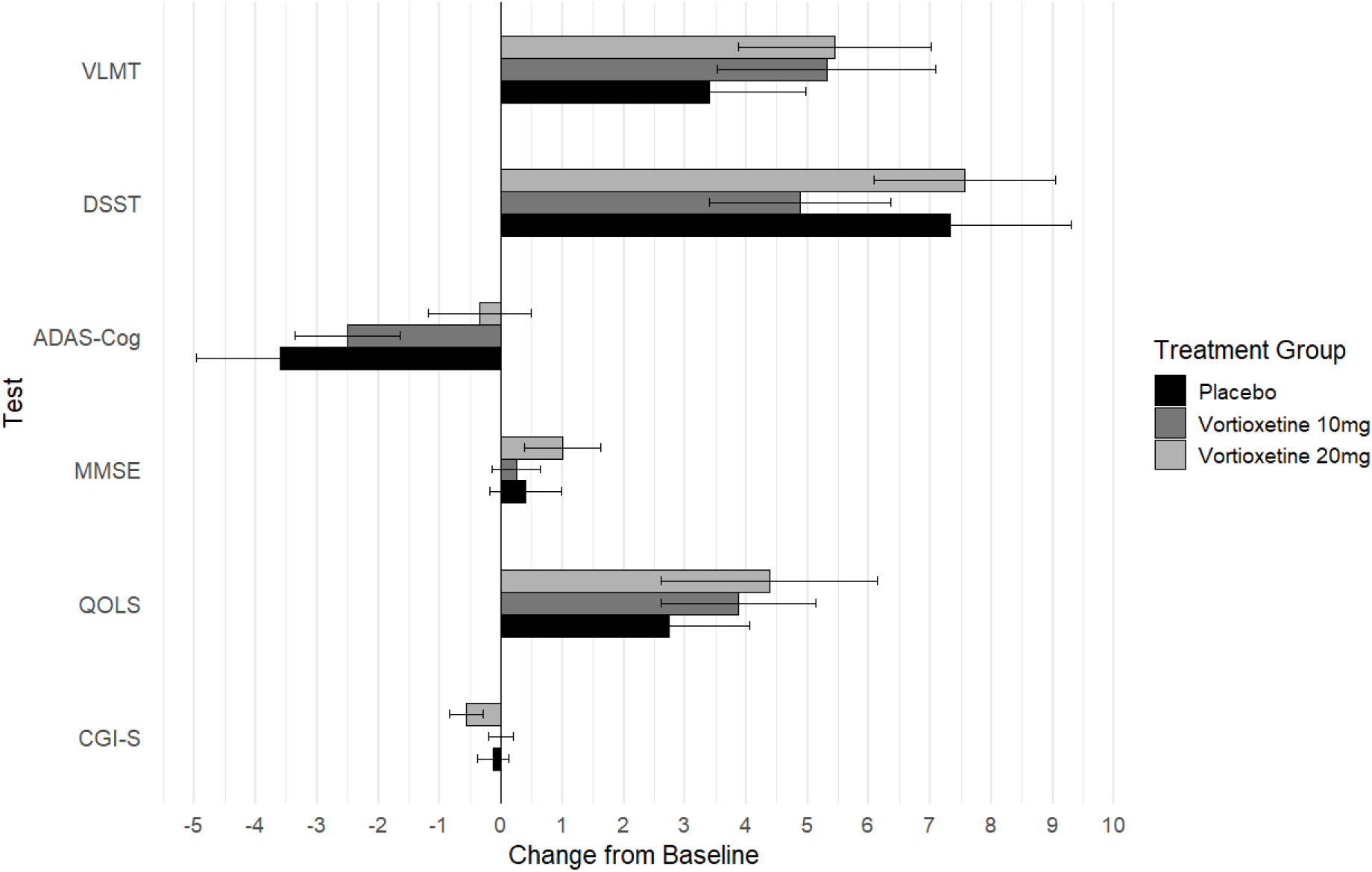
Mean VLMT, DSST, ADAS-Cog, MMSE, QOLS and CGI-S score change from baseline to week 12. Error bars represent 95% confidence intervals (CI) of mean.

**Figure 3.**
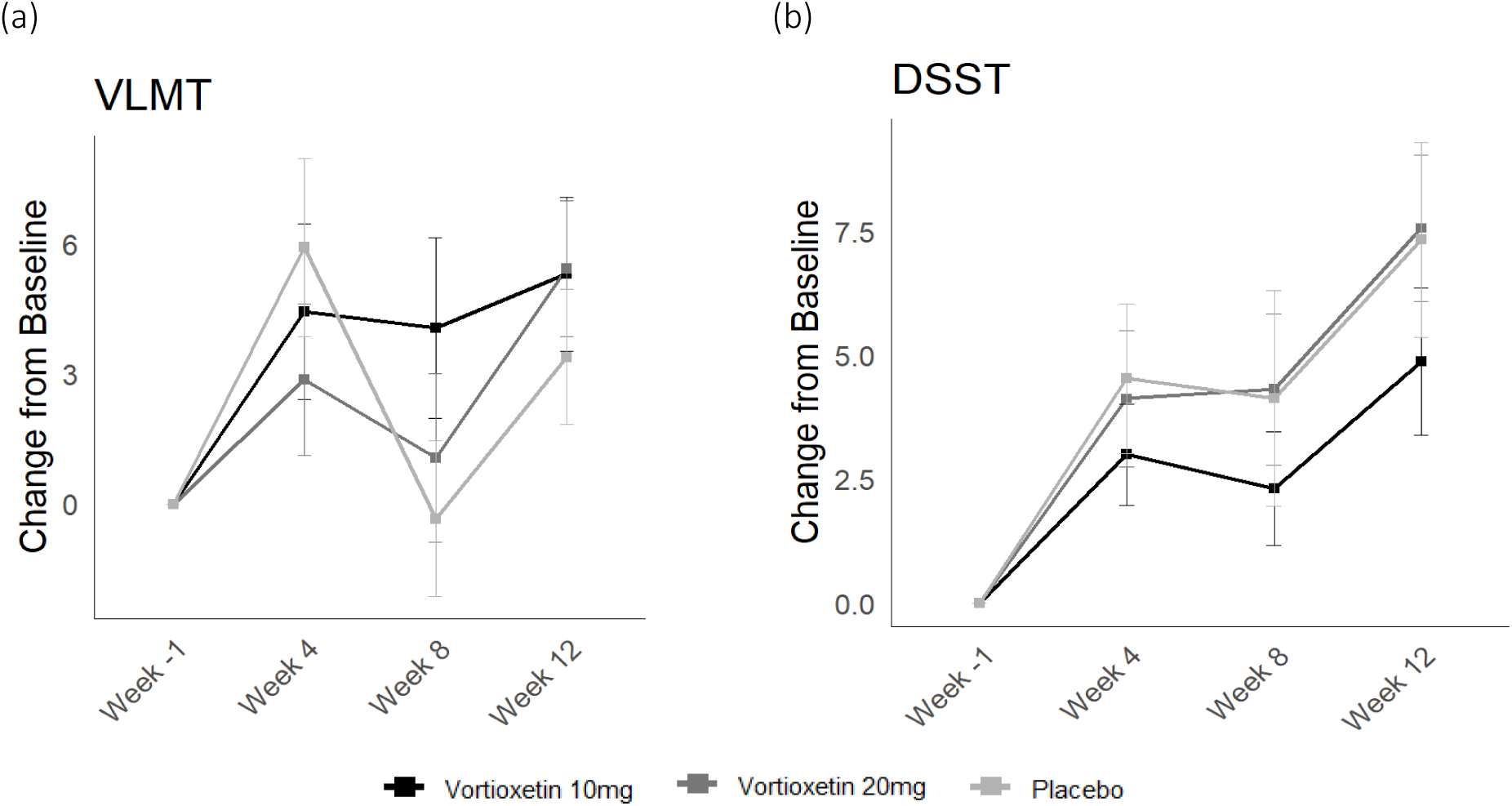
Mean Score Change over Time. (a) Mean VLMT scores at baseline, week 4, week 8 and week 12. (b) Mean DSST scores at baseline, week 4 and week 8.

**Figure 4.**
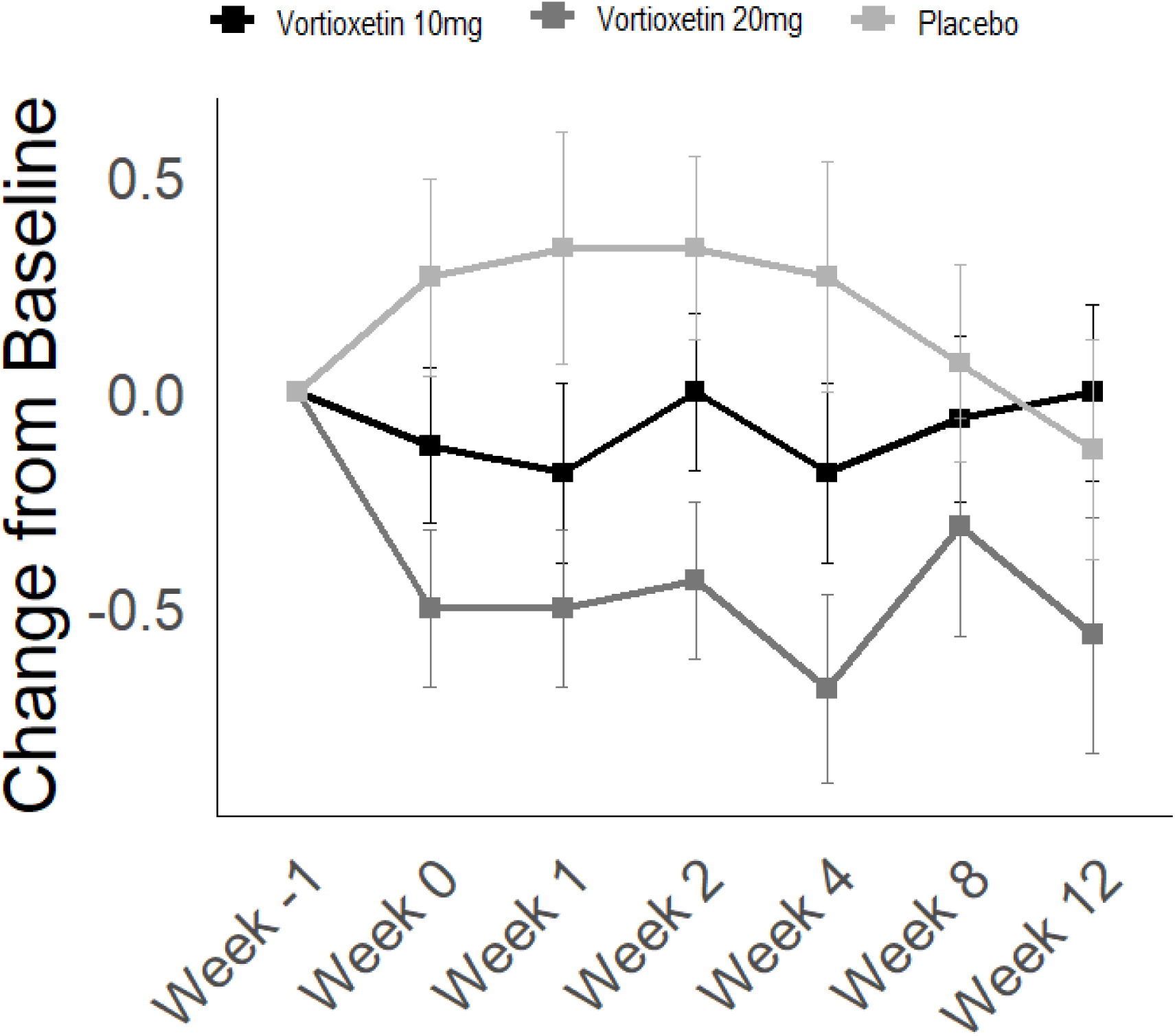
CGI-S Score Change from Baseline. Mean Clinical Global Impression of Severity (CGI-S) at baseline, week 0, week 1, week 2, week 4, week 8 and week 12.

### Safety

Side effects were recorded at weeks 1, 2, 4, 8 and 12, with a mean side effect rating at each visit below 1.4 (UKU scale; Lingjaerde et al, 1987), indicating no or only mild side effects during the trial (Table 4). The most common side effect reported was nausea (25.7% in the 20 mg vortioxetine group with a mean rating of 0.42 and 11.9% in the 10 mg group with a mean rating of 0.16), followed by erectile dysfunction in the 20 mg group (8.6%, mean rating 0.09) and increased dream activity in the 10 mg group (9%, mean rating 0.1).

**Table 4.**
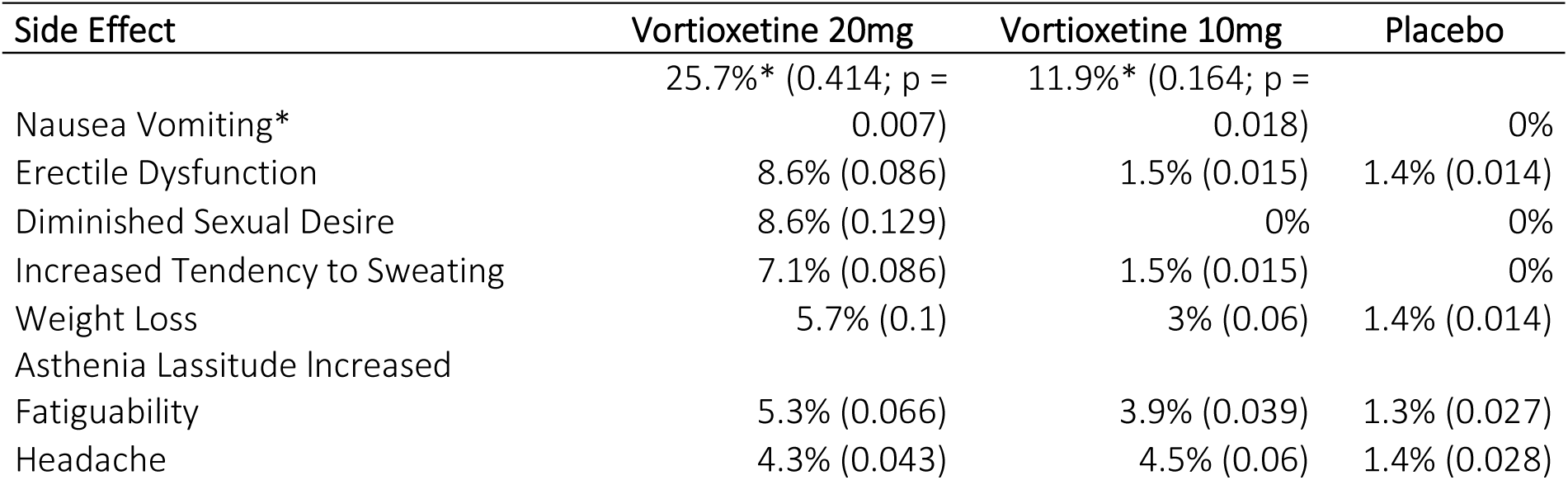
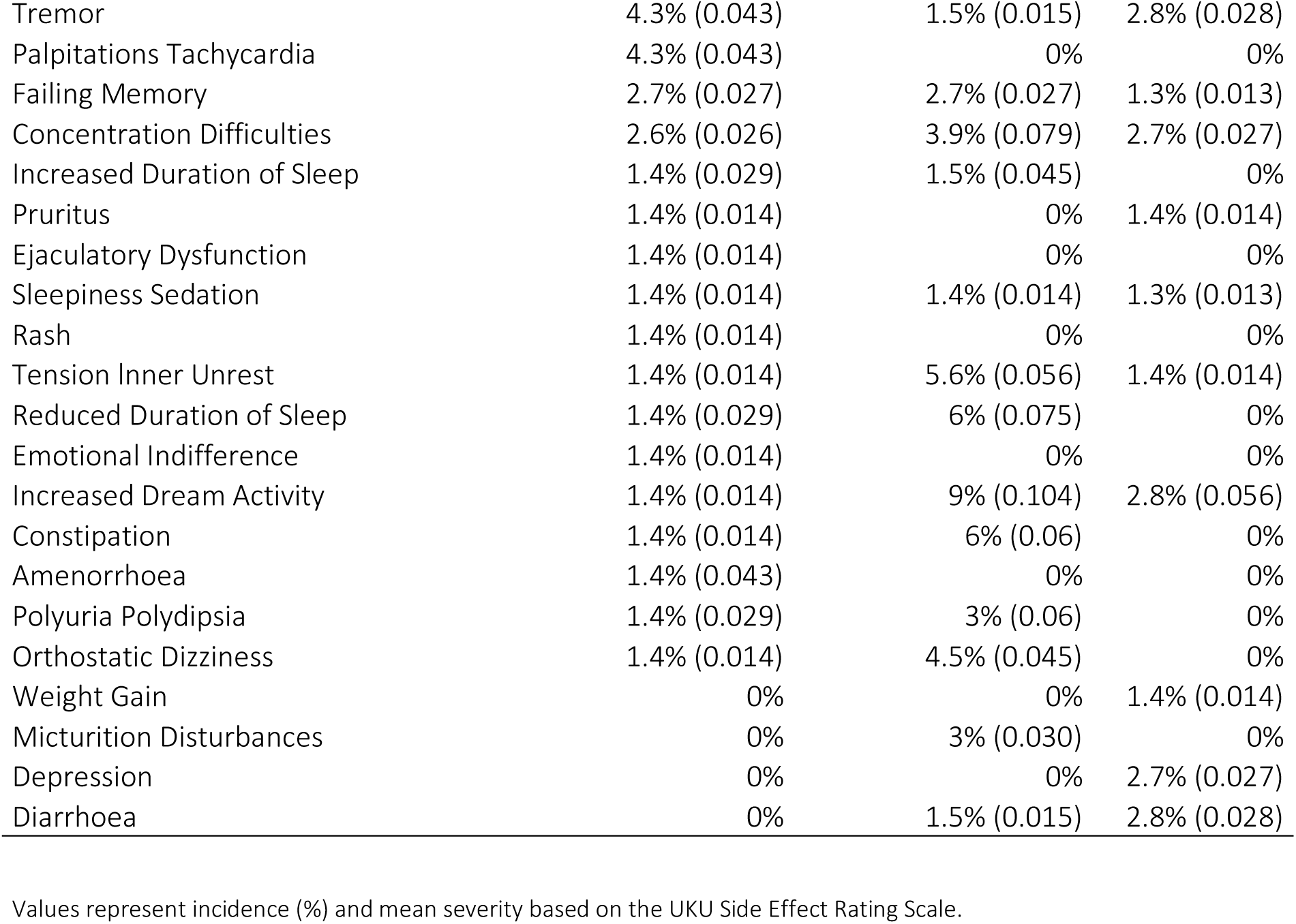
Side Effect Incidence.

## DISCUSSION

The present study assessed the effects of two doses of vortioxetine vs. placebo on cognitive functioning in MCI patients, as measured by several neuropsychological tests. Treatment with vortioxetine was associated with improvements in verbal short-and long-term memory, potentially alleviating cognitive symptoms in neurodegenerative diseases. However, when compared to placebo, no significant group differences could be found.

In a recent meta-analysis, memory, among other cognitive domains, was found to deteriorate prematurely in MCI patients (Chehrehnegar et al., 2020). Thus, the VLMT (“Verbaler Lern-und Merkfähigkeitstest”; Helmstaedter et al, 2001), a German neuropsychological assessment of memory functioning, was used in this study to evaluate this domain in MCI patients. Clinical data validating this test have shown that it is suitable for measuring short-and long-term memory, as well as working memory and verbal recognition, explaining 77% of variance (Helmstaedter et al, 2001), making the test a well-suited outcome measure in this study. In the present study, compared with baseline, the mean VLMT score was significantly improved after 12 weeks of treatment with vortioxetine compared to placebo, indicating a potential treatment effect on verbal learning and memory processes. In contrast, a significant improvement of test scores in the placebo group was observed after 4 weeks, accompanied by a significant decrease in VLMT scores in the following weeks (week 4 to week 8), potentially pointing towards the attenuation of an initially observed placebo effect. These findings are in line with other RCTs comparing vortioxetine and placebo, where significant improvements on verbal learning and memory were observed in vortioxetine treatment in MDD patients (Katona et al., 2012; McIntyre et al., 2014). However, these effects could not be observed at a group level when comparing vortioxetine to placebo, limiting the interpretative value of these findings. Additionally, MMSE scores improved significantly in the 20 mg vortioxetine group after 8 weeks of treatment, whereas scores in the placebo group were stable over time.

Nevertheless, MMSE performance can vary as it is a screening tool broadly assessing several cognitive domains with no clear cut-off scores indicating clinically relevant cognitive impairments for a diagnosis of MCI (Folstein et al, 1983). Thus, it remains unclear if meaningful improvements in cognitive functioning can accurately be detected using the MMSE.

Contrary, significant improvements in DSST performance from baseline were observed in all three groups, with no group differences between placebo and vortioxetine. However, recent studies on the effects of vortioxetine in patients with major depressive disorder (MDD) were able to show significant improvements in DSST performance compared to placebo (Mahableshwarkar et al., 2015; Katona et al, 2012), which could not be replicated in the present study on patients with MCI without depression. These discrepancies might be due to the low specificity in the DSST to determine cognitive function across different cognitive domains, making it a suitable tool to monitor and assess cognitive functioning over time in general, albeit not considering the mainly affected cognitive domain in MCI (Jaeger, 2018). The DSST has been used and deemed robust at predicting outcomes in patients with severe mental illness by measuring a variety of cognitive functions and domains (Dickinson et al., 2007), possibly suggesting that it is suitable in detecting and predicting secondary cognitive deficits, caused by underlying mental illnesses like MDD. This might explain the significant findings in DSST performance in various studies on the effects of vortioxetine in MDD (Katona et al., 2012; McIntyre et al., 2014; Mahableshwarkar et al., 2015). In a recent open-label, single-arm study in patients with MCI, significant improvements in DSST performance were observed when treated with vortioxetine, with a mean score improvement of 7.8 points from baseline to month 3 (Tan & Tan, 2021). As described in the study by Tan & Tan (2021), the score improvement was in line with improvements observed in several RCTs of vortioxetine treatment in MDD (2.8 – 9.0 points at week 8 with a treatment range of 5-20 mg of vortioxetine) (Katona et al., 2012; McIntyre et al., 2014; Mahableshwarkar et al., 2015) and is also in line with the findings of this current study (mean increase of 6 points for the 10mg vortioxetine group, 7.9 points for the 20mg vortioxetine group, and 8.4 points for the placebo group from baseline to week 12). Tan & Tan (2021) attributed these findings to the direct effects of vortioxetine on cognition, rather than to improvements in cognitive functioning through reductions in depressive symptoms. However, the results of the present study indicate that improvements in DSST performance in MCI patients might be independent of vortioxetine treatment. One possible explanation for the increase in DSST scores might be practice effects, which have been linked to a 2.4-DSST-point increase at a 2–3-week retest interval (Jutten et al., 2018). Additionally, cognitive impairment in MCI seems to be highly heterogeneous, with deficits ranging from attention difficulties to impairment in memory, processing speed, and verbal fluency (Chehrehnegar et al., 2020). It seems to be unclear which specific cognitive domain mediates performance on the DSST, thus, the accurate detection of cognitive improvement in patients with MCI might be difficult to achieve with symbol coding tests such as the DSST (Clarke et al, 2012; Zihl et al, 2014).

Similarly, as in DSST test performance scores, ADAS-Cog test performance significantly increased over time in both, the vortioxetine 10 mg and the placebo groups (10 mg: p = 0.02; placebo: p = 0.002).

The ADAS-Cog is a tool used to evaluate the progression of cognitive decline in AD, measuring orientation, memory, executive function and verbal cognition (Rosen et al, 1984). However, according to Kueper et al. (2018) ADAS-Cog scores might be an insufficient assessment in pre-dementia populations, as it may miss changes in cognitive performance and thus, potential treatment benefits, in patients with MCI.

In addition to the assessment of cognitive functioning, as measured by neuropsychological tests during this trial, the subjective well-being of the patients was assessed through questionnaires by the patients and their clinicians. QOLS score improvements were significant only in the 10 mg vortioxetine group, suggesting a subjective improvement in life quality over the course of the study in patients receiving the lower vortioxetine dose, compared with no significant improvements in the other two conditions. This might indicate a dose-dependent effect on subjective well-being, possibly resulting from the treatment-induced improvements in cognitive function, combined with no or only mild side effects when administering the lower vortioxetine dose.

Additionally, clinician-based changes in disease severity and cognitive function improvements were assessed through the Clinical Global Impression Scale (CGI), a tool well suited to evaluate drug efficacy when compared to other, well-known drug efficacy scales used across different psychiatric conditions, such as MDD, Anxiety disorders, or psychotic disorders (Busner & Targum, 2007). A significant group difference between the treatment with 20 mg vortioxetine and placebo was observed in the severity of illness scale (CGI-S), which ranges from 1 = “normal, not ill” to 7 = “severely ill” and is based on the physician’s clinical experience with patients suffering from the same disease. Mean ratings at baseline ranged from 2.6 to 3, indicating patients to be “slightly ill” on average. Scores in the 20mg vortioxetine group dropped from a mean score of 2.75 to a mean score of 2.19 from baseline to week 12, with a score of 2 indicating “borderline psychiatric illness”, thus almost reaching a score describing a healthy population. These findings are in line with clinician ratings carried out in the study by Tan & Tan (2021), where improvements in disease severity changed from very mild impairment to normal status when treated with vortioxetine. In addition, the current study could show that mean placebo scores remained stable over time, with a slight increase in mean severity ratings from baseline to week 8, indicating a clinician-based observation of different disease courses depending on treatment with vortioxetine vs placebo. The clinical global impression of improvement (CGI-I) showed a significant effect of time in the 10 mg vortioxetine group, with a mean score change of 0.6 points from week 1 to week 12. In the first week, average improvement was rated at around 3.9 points in each group, indicating no change. At the end of the trial (week 12) the average improvement rating in the 10 mg vortioxetine group was scored at 3.3 points, pointing toward a minimally improved clinical presentation of illness. This could also be shown by the efficacy index (CGI-EI), as ratings over time reached significance only in the 10 mg vortioxetine group, suggesting a positive effect of 10 mg of vortioxetine on clinician-based symptom changes in MCI patients, combined with well tolerable side effects. As MCI is a rather mild disease, with little to no functional impairments, even subtle changes in clinician ratings as observed in this study could provide important information on treatment success.

Taken together, the results of this RCT suggest an association of vortioxetine treatment with improvements in verbal memory functioning, as seen by the significant time effect of vortioxetine on VLMT performance over the course of 12 weeks. These findings are in line with the cognitive effects of this treatment reported in the literature for patients with MDD (Harrison et al., 2016; Mahableshwarkar et al., 2015) and for patients with MCI (Tan & Tan, 2021). However, the effects of vortioxetine on other domains of cognitive functioning, such as attention or executive function, as described by Harrison et al. (2016) could not be replicated in this study. Several factors might have contributed to this discrepancy, including the relatively small sample size in each treatment arm, as well as the patient population, consisting of only MCI patients without depressive symptoms in the current study, compared to MDD patients in past research. This suggests a specific mechanism of action of vortioxetine on MCI solely targeting memory processes. Another possible explanation might be that improvements in MCI are most sensitively detected through verbal memory tests, thus other measures of cognitive functioning might miss important treatment effects in patients with MCI. A systematic review on the early detection of cognitive disturbances in mild cognitive impairment has found that episodic memory was the best suited predictor of cognitive decline in patients with MCI, with neuropsychological measures such as the RAVLT (Rey Auditory Verbal Learning Test; Rey, 1958) being a reliable detector of early changes in cognitive functioning (Chehrehnegar et al, 2020).

Although objective, performance-based measures are typically used to evaluate cognitive functioning, questionnaires assessing the subjective experience of cognitive decline and the general well-being of patients are crucial for determining the clinical meaningfulness of a specific treatment (Keefe et al., 2013a; McGough & Faraone, 2009). However, discrepancies between the objective and subjective evaluation of cognitive functioning and possible treatment effects have previously been reported and highlight the difficulties in accurately assessing patients’ well-being, cognition and treatment needs (Mahableshwarkar et al., 2015). Specifically, when evaluating patients with subclinical symptomology or mild disease forms such as MCI, subjective patient and clinician ratings can provide meaningful insight into treatment success and current disease state. Objective neuropsychological tests might miss subtle changes in cognitive functioning, as most cognitive assessments were developed to aid in diagnosing more severe forms of cognitive impairment, as observed in AD or other forms of dementia. In accordance with previous clinical studies using vortioxetine as a study medication, the safety profile of vortioxetine treatment showed little to no adverse events, with nausea being the most commonly experienced side effect, as reported in the literature (Baldwin et al., 2016; Tan & Tan, 2021).

This clinical study has several limitations: First, because of the heterogeneous cognitive profile of patients with MCI, choosing appropriate measures to adequately detect cognitive changes in MCI is difficult. In the literature, there is no clear recommendation for cognitive tasks suitable for the diagnosis and evaluation of MCI symptoms, as a consistent diagnostic path for MCI is lacking.

Furthermore, the neuropsychological tests used to quantify cognitive functioning show different criteria for cut-off scores, increasing the risk for false interpretations of test results (Chehrehnegar et al, 2020). Finally, the significant cognitive changes observed in this study were mainly time-based effects rather than treatment group differences, limiting their interpretative value. One possible explanation for this might be the fact that on average, the study subjects in this trial were in the cognitive normal range of the selected neuropsychological tests, leaving only little room for improvement and thus, limiting the possible differences between the placebo and the verum groups. One example for this is the results of the VLMT, with average scores in each treatment group and at each time point above 41 points. According to the VLMT test manual, the average score of a population above 50 years of age is 47.69 points with a SD of 9.93 (Helmstaedter et al, 2001), matching averages in our study population (45.86 ± 11.84). Moreover, the T values described for healthy cognitive functioning in adults above 50 years of age is set with a raw score of 35 (T > 40; Helmstaedter et al., 2001), meaning that in the current study, all participants exhibited a normal range of cognitive functioning in the VLMT. The only measure resulting in a significant time x group difference was the CGI-S scale, with a significant difference in clinician’s severity ratings between patients treated with 20 mg of vortioxetine and those who received placebo. This might underline the difficulty of accurately assessing subtle changes in cognitive functioning with the available neuropsychological tools and thus highlighting the importance of patients’ and clinicians’ evaluation of disease state and cognitive functioning. Therefore, this study also sheds light on the need for reliable diagnostic tools to accurately assess MCI and mild cognitive decline.

## CONCLUSIONS

In this randomized, placebo-controlled, clinical trial, vortioxetine was associated with a potential treatment effect on verbal learning and memory processes, providing possible evidence as a novel pharmacological treatment for MCI and AD patients. Nevertheless, not all outcome measures showed a consistent effect of vortioxetine compared to placebo, potentially suggesting a specific mechanism of action targeted solely at verbal memory. Additionally, effects were observed only over time, rather than between treatment groups, limiting the interpretive value of these findings; thus, further research is needed to clarify the role of vortioxetine in MCI and AD treatment.

## DISCLOSURES

This research was funded by the Austrian Science Fund (FWF) [grant DOI: 10.55776/KLI827, PI: R. Edda Winkler-Pjrek]. For open access purposes, the author has applied a CC BY public copyright license to any author accepted manuscript version arising from this submission.

D. Winkler received lecture fees / authorship honoraria from Angelini, Bristol Myers Squibb, Eli Lilly, Idorsia, Lundbeck, MedAhead, and MedMedia Verlag.

R. Lanzenberger received investigator-initiated research funding from Siemens Healthcare regarding clinical research using PET/MR and travel grants and/or conference speaker honoraria from Janssen-Cilag Pharma GmbH in 2023, and Bruker BioSpin, Shire, AstraZeneca, Lundbeck A/S, Dr. Willmar Schwabe GmbH, Orphan Pharmaceuticals AG, Janssen-Cilag Pharma GmbH, Heel and Roche Austria GmbH., and Janssen-Cilag Pharma GmbH in the years before 2020. He is a shareholder of the start-up company BM Health GmbH, Austria since 2019.

G. Dörl is a recipient of a DOC fellowship of the Austrian Academy of Sciences at the Department of Psychiatry and Psychotherapy, Medical University of Vienna

All other author(s) declare no potential conflicts of interest with respect to the research, authorship, and/or publication of this article.

## Data Availability

The data underlying the analyses in the present study will be made available upon reasonable request to the corresponding author after publication.

## ACKNOWLEDGMENTS

We would like to thank Carina Bum and Victoria Schöber for their work as study physicians.

## Notes

### Clinical Trial

EudraCT 2019-001836-69

### Funding Statement

This research was funded in whole or in part by the Austrian Science Fund (FWF) [grant DOI: 10.55776/KLI827, PI: Edda Winkler-Pjrek; grant DOI: 10.55776/KLI899PI: Dietmar Winkler; grant DOI: 10.55776/PAT6608924, PI: Rupert Lanzenberger; grant DOI: 10.55776/KLI1006, PI: Rupert Lanzenberger]. For open access purposes, the author has applied a CC BY public copyright license to any author accepted manuscript version arising from this submission. Doerl G is a recipient of a DOC fellowship of the Austrian Academy of Sciences at the Department of Psychiatry and Psychotherapy, Medical University of Vienna.

### Author Declarations

Ethics Committee of the Medical University of Vienna gave ethical approval for this work

## REFERENCES

Albert, M. S., DeKosky, S. T., Dickson, D., Dubois, B., Feldman, H. H., Fox, N. C., Gamst, A., Holtzman, D. M., Jagust, W. J., Petersen, R. C., Snyder, P. J., Carrillo, M. C., Thies, B., & Phelps, C. H. (2011). The diagnosis of mild cognitive impairment due to Alzheimer’s disease: Recommendations from the National Institute on Aging–Alzheimer’s Association workgroups on diagnostic guidelines for Alzheimer’s disease. Alzheimer’s & Dementia, 7, 270–279. 10.1016/j.jalz.2011.03.008

Alenina, N., & Klempin, F. (2015). The role of serotonin in adult hippocampal neurogenesis. Behavioural Brain Research, 277, 49–57. 10.1016/j.bbr.2014.07.038

Baldwin, D. S., Chrones, L., Florea, I., Nielsen, R., Nomikos, G. G., Palo, W., & Reines, E. (2016). The safety and tolerability of vortioxetine: Analysis of data from randomized placebo-controlled trials and open-label extension studies. Journal of Psychopharmacology, 30, 242–252. 10.1177/0269881115622561

Barrett, F. S., Workman, C. I., Sair, H. I., Savonenko, A. V., Kraut, M. A., Sodums, D. J., Joo, J. H., Nassery, N., Marano, C. M., Munro, C. A., Brandt, J., Zhou, Y., Wong, D. F., & Smith, G. S. (2017). Association between serotonin denervation and resting-state functional connectivity in mild cognitive impairment. Human Brain Mapping, 38, 551–560. 10.1002/hbm.23399

Baune, B. T., Brignone, M., & Larsen, K. G. (2018). A network meta-analysis comparing effects of various antidepressant classes on the Digit Symbol Substitution Test (DSST) in major depressive disorder. International Journal of Neuropsychopharmacology, 21, 97–107. 10.1093/ijnp/pyx072

Callahan, C. M. (2017). Alzheimer’s disease: Individuals, dyads, communities, and costs. Journal of the American Geriatrics Society, 65, 892–895. 10.1111/jgs.14736

Chehrehnegar, N., Nejati, V., Shati, M., Rashedi, V., Lotfi, M., Adelirad, F., & Foroughan, M. (2020). Early detection of cognitive disturbances in mild cognitive impairment: A systematic review of observational studies. Psychogeriatrics, 20, 212–228. 10.1111/psyg.12484

Chen, J., Xiang, P., Duro-Castano, A. et al. (2025). Rapid amyloid-β clearance and cognitive recovery through multivalent modulation of blood–brain barrier transport. Sig Transduct Target Ther 10, 331. 10.1038/s41392-025-02426-1

Clarke, P. J., Ailshire, J. A., House, J. S., Morenoff, J. D., King, K., & Melendez, R. (2012). Cognitive function in the community setting: The neighbourhood as a source of cognitive reserve? Journal of Epidemiology & Community Health, 66, 730–736. 10.1136/jech.2010.115477

D’Agostino, A., English, C. D., & Rey, J. A. (2015). Vortioxetine (Brintellix): A new serotonergic antidepressant. P & T, 40, 36–40.

Damoiseaux, J. S., Prater, K. E., Miller, B. L., & Greicius, M. D. (2012). Functional connectivity tracks clinical deterioration in Alzheimer’s disease. Neurobiology of Aging, 33, 828.e19–828.e30. 10.1016/j.neurobiolaging.2011.06.024

Devita, M., Masina, F., Mapelli, D., Anselmi, P., Sergi, G., & Coin, A. (2021). Acetylcholinesterase inhibitors and cognitive stimulation in mild Alzheimer’s disease: Combined and alone. Aging Clinical and Experimental Research, 33, 3039–3045. 10.1007/s40520-021-01892-5

Dickinson, D., Ramsey, M. E., & Gold, J. M. (2007). Overlooking the obvious: A meta-analytic comparison of digit symbol coding tasks and other cognitive measures in schizophrenia. Archives of General Psychiatry, 64, 532–542. 10.1001/archpsyc.64.5.532

Dou, K. X., Tan, M. S., Tan, C. C., et al. (2018). Comparative safety and effectiveness of cholinesterase inhibitors and memantine for Alzheimer’s disease: A network meta-analysis of 41 randomized controlled trials. Alzheimer’s Research & Therapy, 10, 126. 10.1186/s13195-018-0457-9.

Eickhoff, S.B., Constable, R.T., Yeo, B.T.T. (2018). Topographic organization of the cerebral cortex and brain cartography. Neuroimage; 170: 332–347.

First, M., Williams, J., Karg, R., & Spitzer, R. (2015). Structured Clinical Interview for DSM-5 – Research Version (SCID-5 for DSM-5, Research Version; SCID-5-RV). Arlington, VA: American Psychiatric Association.

Folstein, M. F., Robins, L. N., & Helzer, J. E. (1983). The Mini-Mental State Examination. Archives of General Psychiatry, 40, 812. 10.1001/archpsyc.1983.01790060092014

Fox, N. C., Belder, C., Ballard, C., et al. (2025). Treatment for Alzheimer’s disease. The Lancet. Advance online publication. S0140-6736(25)01329-7

Gauthier, S., Feldman, H., Hecker, J., Vellas, B., Emir, B., & Subbiah, P. (2002). Functional, cognitive and behavioral effects of donepezil in patients with moderate Alzheimer’s disease. Current Medical Research and Opinion, 18, 347–354. 10.1185/030079902125001716

Grundman, M., Petersen, R. C., Ferris, S. H., Thomas, R. G., Aisen, P. S., Bennett, D. A., Foster, N. L., Jack, C. R., Jr., Galasko, D. R., Doody, R., Kaye, J., Sano, M., Mohs, R., Gauthier, S., Kim, H. T., Jin, S., Schultz, A. N., Schafer, K., Mulnard, R., van Dyck, C. H., Mintzer, J., Zamrini, E. Y., Cahn-Weiner, D., & Thal, L. J. (2004). Mild cognitive impairment can be distinguished from Alzheimer disease and normal aging for clinical trials. Archives of Neurology, 61, 59–66. 10.1001/archneur.61.1.59

Harrison, J.E., Lophaven, S., Olsen, C.K. (216). Which Cognitive Domains are Improved by Treatment with Vortioxetine? Int J Neuropsychopharmacol.

Hasselbalch, S. G., Madsen, K., Svarer, C., Pinborg, L. H., Holm, S., Paulson, O. B., Waldemar, G., & Knudsen, G. M. (2008). Reduced 5-HT₂A receptor binding in patients with mild cognitive impairment. Neurobiology of Aging, 29, 1830–1838. 10.1016/j.neurobiolaging.2007.04.015

Helmstaedter, C., Lendt, M., & Lux, S. (2001). Verbaler Lern-und Merkfähigkeitstest (1. Aufl.). Göttingen: Beltz Test.

Jaeger, J. (2018). Digit Symbol Substitution Test: The case for sensitivity over specificity in neuropsychological testing. Journal of Clinical Psychopharmacology, 38(5), 513–519. 10.1097/JCP.0000000000000941

Katona, C., Hansen, T., & Olsen, C. K. (2012). A randomized, double-blind, placebo-controlled, duloxetine-referenced, fixed-dose study comparing the efficacy and safety of Lu AA21004 in elderly patients with major depressive disorder. International Clinical Psychopharmacology, 27(4), 215–223. 10.1097/YIC.0b013e3283542457

Kueper, J. K., Speechley, M., & Montero-Odasso, M. (2018). The Alzheimer’s Disease Assessment Scale-Cognitive Subscale (ADAS-Cog): Modifications and responsiveness in pre-dementia populations — A narrative review. Journal of Alzheimer’s Disease, 63(2), 423–444. 10.3233/JAD-170991

Lanctôt, K. L., Hussey, D. F., Herrmann, N., Black, S. E., Rusjan, P. M., Wilson, A. A., Houle, S., Kozloff, N., Verhoeff, N. P., & Kapur, S. (2007). A positron emission tomography study of 5-hydroxytryptamin-1A receptors in Alzheimer disease. American Journal of Geriatric Psychiatry, 15, 888–898. 10.1097/01.JGP.0000256853.42022.12

Lezak, M. D., Howieson, D. B., Bigler, E. D., & Tranel, D. (2012). Neuropsychological Assessment (5^th^ ed.). New York, NY: Oxford University Press.

Langa, K. M., & Levine, D. A. (2014). The diagnosis and management of mild cognitive impairment: A clinical review. JAMA, 312, 2551–2561. 10.1001/jama.2014.13806

Lingjaerde, O., Ahlfors, U. G., Bech, P., Dencker, S. J., & Elgen, K. (1987). The UKU side effect rating scale: A comprehensive rating scale for psychotropic drugs and a cross-sectional study of side effects in neuroleptic-treated patients. Acta Psychiatrica Scandinavica. Supplementum, 334, 1–100.

Mahableshwarkar, A. R., Zajecka, J., Jacobson, W., Chen, Y., & Keefe, R. S. (2015). A randomized, placebo-controlled, active-reference, double-blind, flexible-dose study of the efficacy of vortioxetine on cognitive function in major depressive disorder. Neuropsychopharmacology, 40, 2025–2037. 10.1038/npp.2015.52

McGough, J. J., & Faraone, S. V. (2009). Estimating the size of treatment effects: Moving beyond p values. Psychiatry (Edgmont), 6, 21–29.

McIntyre, R. S., Florea, I., Tonnoir, B., Loft, H., Lam, R. W., & Christensen, M. C. (2017). Efficacy of vortioxetine on cognitive functioning in working patients with major depressive disorder. Journal of Clinical Psychiatry, 78, 115–121. 10.4088/JCP.16m10756

McIntyre, R. S., Lophaven, S., & Olsen, C. K. (2014). A randomized, double-blind, placebo-controlled study of vortioxetine on cognitive function in depressed adults. International Journal of Neuropsychopharmacology, 17(10), 1557–1567. 10.1017/S1461145714000546

Mork, A., Pehrson, A., Brennum, L. T., Nielsen, S. M., Zhong, H., Lassen, A. B., Miller, S., Westrich, L., Boyle, N. J., Sanchez, C., Fischer, C. W., Liebenberg, N., Wegener, G., Bundgaard, C., Hogg, S., Bang-Andersen, B., & Stensbøl, T. B. (2012). Pharmacological effects of Lu AA21004: A novel multimodal compound for the treatment of major depressive disorder. Journal of Pharmacology and Experimental Therapeutics, 340, 666–675. 10.1124/jpet.111.190882

Nelson, R. L., Guo, Z., Halagappa, V. M., Pearson, M., Gray, A. J., Matsuoka, Y., Brown, M., Martin, B., Iyun, T., Maudsley, S., Clark, R. F., & Mattson, M. P. (2007). Prophylactic treatment with paroxetine ameliorates behavioral deficits and retards the development of amyloid and tau pathologies in 3×TgAD mice. Experimental Neurology, 205, 166–176. 10.1016/j.expneurol.2006.12.012

Offenbacher, M., Sauer, S., Kohls, N., Waltz, M., & Schoeps, P. (2012). Quality of life in patients with fibromyalgia: Validation and psychometric properties of the German Quality of Life Scale (QOLS-G). Rheumatology International, 32, 3243–3252. 10.1007/s00296-011-2262-5

Pae, C.U, Wang, S.M., Han, C., Lee, S.J., Patkar, A.A., Masand, P.S., Serretti, A. (2015).Vortioxetine: a meta-analysis of 12 short-term, randomized, placebo-controlled clinical trials for the treatment of major depressive disorder. J Psychiatry Neurosci, 40, 174–186.

Petersen, R. C. (2004). Mild cognitive impairment as a diagnostic entity. Journal of Internal Medicine, 256, 183–194. 10.1111/j.1365-2796.2004.01388.x

Petersen, R. C. (2016). Mild cognitive impairment. *Continuum (Minneapolis*, Minn.), 22, 404–418. 10.1212/CON.0000000000000313

Petersen, R. C., Smith, G. E., Waring, S. C., Ivnik, R. J., Tangalos, E. G., & Kokmen, E. (1999). Mild cognitive impairment: Clinical characterization and outcome. Archives of Neurology, 56, 303–308. 10.1001/archneur.56.3.303

Rey, A. (1958). Rey Auditory Verbal Learning Test (RAVLT) [Database record]. APA PsycTests. 10.1037/t09066-000

Rosen, W. G., Mohs, R. C., & Davis, K. L. (1984). A new rating scale for Alzheimer’s disease. The American Journal of Psychiatry, 141, 1356–1364. 10.1176/ajp.141.11.1356

Sanchez, C., Asin, K. E., & Artigas, F. (2015). Vortioxetine, a novel antidepressant with multimodal activity: review of preclinical and clinical data. Pharmacology & Therapeutics, 145, 43–57. 10.1016/j.pharmthera.2014.09.006

Schneider, L. S., Dagerman, K. S., Higgins, J. P., & McShane, R. (2011). Lack of evidence for the efficacy of memantine in mild Alzheimer disease. Archives of Neurology, 68, 991–998. 10.1001/archneurol.2011.74

Sheline, Y. I., & Raichle, M. E. (2013). Resting state functional connectivity in preclinical Alzheimer’s disease. Biological Psychiatry, 74, 340–347. 10.1016/j.biopsych.2013.04.028

Smith, G. S., Barrett, F. S., Joo, J. H., Nassery, N., Savonenko, A., Sodums, D. J., Marano, C. M., Munro, C. A., Brandt, J., Kraut, M. A., Zhou, Y., Wong, D. F., & Workman, C. I. (2017). Molecular imaging of serotonin degeneration in mild cognitive impairment. Neurobiology of Disease, 105, 33–41. 10.1016/j.neurobioldis.2017.05.003

Szeto, J. Y., & Lewis, S. J. (2016). Current treatment options for Alzheimer’s disease and Parkinson’s disease dementia. Current Neuropharmacology, 14, 326–338. 10.2174/1570159X13666151216115503

Tan, S. N., & Tan, C. (2021). Vortioxetine improves cognition in mild cognitive impairment. International Clinical Psychopharmacology, 36(6), 279–287. 10.1097/YIC.0000000000000376

World Health Organization. (2025, November 26). Fact sheet: Dementia. Retrieved from https://www.who.int/news-room/fact-sheets/detail/dementia

Yee, A., Ng, C. G., & Seng, L. H. (2018). Vortioxetine treatment for anxiety disorder: A meta-analysis study. Current Drug Targets, 19, 1412–1423. 10.2174/1389450119666180426110557

Zihl, J., Fink, T., Pargent, F., Ziegler, M., & Bühner, M. (2014). Cognitive reserve in young and old healthy subjects: Differences and similarities in a testing-the-limits paradigm with DSST. PLoS ONE, 9(e84590). 10.1371/journal.pone.0084590

